# Individualized Forecasting of Headache Attack Risk Using a Continuously Updating Model

**DOI:** 10.64898/2026.04.20.26350119

**Authors:** Timothy T. Houle, Adriana Lebowitz, Ivana Chtay, Twinkle Patel, Donald D. McGeary, Dana P. Turner

## Abstract

**Importance:** Migraine attacks often occur unpredictably, limiting the ability of individuals to initiate timely preventive or preemptive treatment. Short-term probabilistic forecasting of migraine risk could enable more targeted management strategies.

**Objective:** To externally validate the previously developed Headache Prediction Model (HAPRED-I), evaluate an updated continuously learning model (HAPRED-II), and assess the feasibility and short-term safety of delivering individualized probabilistic migraine forecasts directly to patients.

**Design, Setting, and Participants:** Prospective 8-week cohort study conducted remotely at two academic medical centers in the United States (Massachusetts General Hospital and Wake Forest Health Sciences) between 2015 and 2019. Adults with recurrent migraine or tension-type headache completed twice-daily electronic diaries. A total of 230 participants contributed 23,335 diary entries across 11,862 participant-days of observation.

**Main Outcomes and Measures:** Occurrence of a headache attack within 24 hours following each evening diary entry. Model performance was evaluated using discrimination (area under the receiver operating characteristic curve [AUC]) and calibration.

**Results:** External validation of HAPRED-I demonstrated modest discrimination (AUC, 0.59; 95% CI, 0.57–0.61) and poor calibration, with predicted probabilities consistently exceeding observed headache risk. In contrast, the continuously updating HAPRED-II model demonstrated progressive improvement in predictive performance as participant-specific data accumulated. Discrimination increased from an AUC of 0.59 (95% CI, 0.57–0.61) during the first 14 days to 0.66 (95% CI, 0.63–0.70) after the first month, accompanied by improved calibration across predicted risk levels. Over the study period, 6999 individualized forecasts were delivered directly to participants. No evidence suggested that receipt of forecasts was associated with increasing headache frequency or worsening predicted headache risk trajectories.

**Conclusions and Relevance:** A static migraine forecasting model demonstrated limited transportability to new individuals. In contrast, models that continuously update within individuals may improve predictive accuracy over time and enable real-time delivery of personalized migraine risk forecasts. Further work incorporating richer physiologic and contextual predictors will likely be necessary before such systems can reliably guide clinical treatment decisions.

## Introduction

The experience of headache is inherently unpredictable.^1^ Individuals can rapidly experience an attack at any time during their day and are often caught unprepared to treat a novel attack.^2^ Although individuals typically have a long list of beliefs about what causes their attacks (i.e., ‘triggers’),^3^ we do not know the causal agent for any given attack; thus, preventing these attacks using behavioral avoidance is unwise and tailoring focused management strategies remains challenging.(see: ^4^) Medication strategies that lower the daily risk for attacks can be effective for many individuals,^5^ but such strategies can be poorly tolerated,^6^ expensive, (see: ^7^) and are often discontinued.^8^ By allowing targeted, ‘preemptory’ treatment based on attack risk (see: ^9–11^), a method to predict the future onset of an attack has the potential to revolutionize the treatment and research of migraine.^10,12^

In 2017 Houle et al. developed, estimated, and internally validated a prognostic model, the Headache Prediction Model (HAPRED-I), designed to estimate the probability of a migraine attack in the next 24 hours.^13^ The model deliberately adopted a parsimonious approach, using only the presence versus absence of a current headache and the total score on the Daily Stress Inventory as predictors. The model showed promising discrimination, with an AUC of 0.73 (95% CI 0.71–0.75) in the derivation sample, though attenuated in leave-one-patient-out internal validation (AUC 0.65, 95% CI 0.60–0.67).^13^ The model was well calibrated but exhibited limited resolution; that is, its predictions clustered near the sample’s baseline headache rate.

Since then, the field has seen several additional migraine-attack forecasting models. Holsteen et al, (2020) explored the use of within-person models based on common triggers (e.g., caffeine, stress, menstruation) in 178 participants and reported only modest discrimination, AUC 0.56 (95%CI: 0.54 to 0.58).^14^ Stubberud et al. (2023) explored machine learning models (e.g., random forest) combining mobile headache-diary entries with simple wearable physiological data (heart rate, skin temperature, muscle tension) in 18 patients and achieved an AUC of 0.62 in a hold-out partition.^15^Tsai et al. (2025) implemented individualized machine learning models using digital headache-diary data in 25 participants and reported a mean AUC 0.83 for predicting a next-day attack,^16^ though this discrimination is likely overly-optimistic given the methods used and lack of a validation sample.

There were three primary objectives of this investigation. First, we sought to externally validate the previously developed Headache Prediction Model (HAPRED-I) in an independent sample of individuals with recurrent episodic migraine. Second, we developed and evaluated an updated model, HAPRED-II, which extends the original specification by incorporating continuous Bayesian updating of individual-level parameters, allowing probabilistic forecasts to adapt dynamically as new daily data are accumulated. A Bayesian approach is uniquely suited to allow updating while learning individual differences of model parameters.^19^ Third, we examined the potential clinical implications of delivering these individualized forecasts directly to participants at the end of each day, including whether receiving such predictions influenced headache outcomes.

## Methods

### Study Design and Participants

This was a prospective, 8-week longitudinal cohort study designed for the external validation and Bayesian extension of the previously developed HAPRED-I model for forecasting daily migraine onset risk (now termed HAPRED-II). Participants were recruited from two academic medical centers: Wake Forest Health Sciences (Winston-Salem, NC) and Massachusetts General Hospital (Boston, MA). Recruitment occurred between 5/20/2015 to 1/30/2016 in Winston-Salem and 3/13/2018 to 10/18/2019 in Boston. All study activities, including screening, consent, data entry, and forecast delivery, were conducted remotely via secure web-based systems. The study was approved by the Institutional Review Boards at both participating institutions. All participants completed electronic informed consent prior to data collection.

Eligibility criteria were designed to closely mirror those used in the original HAPRED-I model development study,^13^ with the intention of broadly selecting community-dwelling individuals with recurrent headache who might reasonably benefit from a headache forecasting system. Adults aged 18 years or older with an International Classification of Headache Disorders, 2nd or 3rd edition (ICHD-3; ^17^) provisional diagnosis of migraine (with or without aura) or tension-type headache assessed using a structured diagnostic interview, and who reported experiencing at least one headache attack per month, were considered for enrollment. Participants were required to have internet access and the ability to complete electronic diaries in English. Individuals were excluded if they had a secondary headache disorder or reported a recent major change in headache symptoms or treatment within the prior six weeks.

### Data Collection

Participants completed enrollment questionnaires using REDCap forms,^18^ and then completed twice-daily electronic diaries (morning and evening) for 8 weeks. Diary entries recorded headache presence and daily stress exposures. After completion of an initial 7- to 14-day ‘warm-up’ period,^19^ individualized probabilistic forecasts estimating the likelihood of experiencing a headache attack within the next 24 hours were delivered following each evening diary entry. The Supplemental Methods displays an example of a delivered forecast.

### Predictor Variables

Consistent with the HAPRED-I model specification, two predictors were used, the degree of ‘daily hassles’ measured by the Daily Stress Inventory (DSI)^20^ and the presence-absence of a current headache attack at the time of the diary entry, so these two variables were used in the model to deliver forecasts in this external validation study.

### Primary Outcome

The primary outcome for model validation was the occurrence of a headache attack within the 24-hour period following each evening diary entry. Headache occurrence was defined as any diary report of head pain greater than zero, recorded in either the subsequent morning or evening diary entry of the next calendar day.

### Prediction Models

Two prediction approaches were evaluated. First, the original HAPRED-I model was externally validated by applying the previously published intercept and regression coefficients without re-estimation. Second, an updated model (HAPRED-II) was implemented that allowed model parameters to update continuously within individuals as longitudinal data accumulated. This approach estimated participant-specific intercepts and predictor effects using Bayesian updating, enabling forecasts to adapt dynamically to each participant’s observed data over time. Additional model specification details are provided in the Supplement.

### Sample Size Considerations

Since the origination of the study, Riley and colleagues, have offered advice on planning for the development (2019)^21^ and external validation (2021)^22^ of prediction models. This advice was examined for the current analysis with the available sample size exceeding the recommended sample size for HAPRED-II evaluation. See Supplemental Methods for full details.

### Statistical Analysis

Model performance was evaluated using discrimination and calibration.

Discrimination was assessed using the area under the receiver operating characteristic curve (AUC). Calibration was examined by comparing predicted probabilities with observed event frequencies using locally weighted smoothing. For the continuously updating HAPRED-II model, predictive performance was evaluated across successive epochs of observation (<14 days, 14–27 days, and >27 days) to examine improvements as participant-specific data accumulated. Confidence intervals for AUC estimates were derived using participant-clustered bootstrap resampling. All analyses were conducted using R statistical software (version 4.5.1). Additional analytic details are provided in the Supplemental Methods.

## Results

Participants were recruited from two clinical sites (Massachusetts General Hospital [MGH], *n* = 62; Wake Forest University [WFU], *n* = 168). A total of 17 individuals were enrolled but did not complete any headache diaries and were therefore excluded from all analyses; Table 1 summarizes the characteristics of the 230 participants included in the analysis. A provisional diagnosis of a migraine headache disorder could be confirmed in 194 of 224 (87%) individuals, with 30 (13%) individuals experiencing tension-type headaches; 6 individuals did not experience sufficient attacks to confirm a diagnosis. The sample was predominantly female at both sites (MGH 90.3%, WFU 90.6%) and largely White (MGH 73.8%, WFU 81.0%), with similar representation of Black, Asian, American Indian/Alaskan Native, and Hispanic participants across sites. Baseline clinical characteristics were generally well balanced between sites, with small standardized mean differences (SMDs) for most variables.

**Table 1.** Sample Characteristics.

|  | MGH<br>N = 62 | WFU<br>N = 168 | SMD |
| --- | --- | --- | --- |
| Sex - Female (%) | 56 (90.3) | 96 (90.6) | 0.080 |
| Race (%) |  |  |  |
| White | 45 (73.8) | 85 (81.0) | 0.364 |
| Black | 8 (13.1) | 13 (12.4) |  |
| Asian | 7 (11.5) | 3 (2.9) |  |
| American Indian or Alaskan Native | 1 (1.6) | 4 (3.8) |  |
| Hispanic (%) | 7 (11.3) | 5 (4.9) | 0.238 |
| Age | 29 [24.0, 39.0] | NA |  |
| Depression Screen (%) | 10 (16.1) | 50 (29.8) | 0.329 |
| Anxiety Screen (%) | 13 (21.0) | 61 (36.3) | 0.344 |
| MIDAS | 22.5 [8.0, 36.0] | 16.62 [7.75, 29.0] | 0.332 |
| Headache Frequency | 15 [10.0, 30.0] | 18 [10.0, 30.0] | 0.039 |
| Pain Intensity (0 to 10) | 6 [5.0, 7.0] | 6 [5.0, 7.0] | 0.158 |
| Headache Triggers |  |  |  |
| Red wine | 2 [0.0, 8.0] | 0 [0.0, 3.50] | 0.372 |
| Other alcoholic beverages | 2.5 [0.0, 5.75] | 1 [0.0, 3.0] | 0.34 |
| MSG | 1 [0.0, 3.0] | 0 [0.0, 3.0] | 0.071 |
| Chocolate | 0 [0.0, 1.0] | 0 [0.0, 0.0] | 0.228 |
| Caffeinated beverages | 0 [0.0, 2.75] | 0 [0.0, 2.0] | 0.18 |
| Eating less while dieting | 5 [2.0, 8.0] | 3 [0.0, 6.0] | 0.381 |
| Delaying a meal | 6 [5.0, 8.0] | 5 [3.0, 7.50] | 0.288 |
| Skipping a meal | 7 [4.0, 8.0] | 5 [3.0, 8.0] | 0.226 |
| Snacking instead of meal | 2 [0.0, 5.0] | 1 [0.0, 3.0] | 0.307 |
| Sleeping or napping more | 1 [0.0, 4.75] | 1 [0.0, 3.0] | 0.17 |
| Sleeping or napping less | 7 [3.0, 8.0] | 4 [2.0, 7.0] | 0.407 |
| Insomnia | 6 [1.0, 8.0] | 3 [0.0, 6.0] | 0.317 |
| Exercising to exhaustion | 1 [0.0, 4.75] | 0 [0.0, 3.0] | 0.249 |
| Feeling overly fatigued | 5 [3.0, 8.0] | 5 [2.0, 7.0] | 0.279 |
| Feeling very pressured | 7.5 [5.0, 9.0] | 6.5 [4.0, 8.0] | 0.234 |
| Feeling very upset | 4.5 [0.25, 7.0] | 3 [1.0, 6.0] | 0.177 |
| Feeling very irritable | 3 [1.0, 7.0] | 3 [1.0, 7.0] | 0.06 |
| Feeling very tense or anxious | 6 [2.0, 8.0] | 5 [2.0, 8.0] | 0.146 |
| Odors | 6 [0.25, 8.0] | 3 [1.0, 6.0] | 0.339 |
| Smoke | 2 [0.0, 7.75] | 2 [0.0, 5.0] | 0.102 |
| Chemical vapors | 3 [0.0, 6.75] | 3 [0.0, 6.0] | 0.034 |
| Strong perfume/cologne | 4 [1.0, 8.0] | 4 [1.0, 8.0] | 0.035 |
| Bright light | 6 [2.25, 8.0] | 3 [1.0, 6.0] | 0.507 |
| Flickering light | 5 [0.25, 8.0] | 3 [1.0, 6.0] | 0.344 |
| Glare | 4 [1.0, 6.75] | 3 [0.25, 6.0] | 0.247 |
| Hot environment | 4 [1.0, 7.0] | 2 [0.0, 6.0] | 0.204 |
| Loud noises | 5 [1.25, 8.0] | 3 [1.0, 6.0] | 0.402 |
| Weather or barometric change | 5 [1.0, 8.0] | 4 [1.0, 8.0] | 0.171 |

Median headache burden was substantial, with MIDAS scores of 22.5 [IQR 8–36] at MGH and 16.6 [7.75–29] at WFU, and frequent headaches reported at both sites (median 5 [3.3–10] vs. 6 [3.3–10] days/30-day month, respectively). Median pain intensity was identical (6/10 at both sites). Screening rates for depression and anxiety were higher at WFU, though differences remained modest. Across both cohorts, participants endorsed a broad range of headache triggers, with highly similar distributions across sites.

### External Validation of HAPRED-I

For the external validation of the original HAPRED-I study, N = 230 individuals were observed over a total of 23,335 AM and PM diary entry occasions on 11,862 participant-days. Non-missing theoretical forecasts could be calculated for 9439 of 11658 (81%) PM entries and evaluated for 8986 of 9439 (95%) of forecasts. A headache attack meeting the primary outcome was observed on 3067 of 8986 (34.1%) of evaluable forecast-days. Discrimination of the HAPRED-I was only modest, with an area under the receiver operating characteristic curve (AUC) of 0.591 (95% CI: 0.574–0.608), indicating performance somewhat better than chance in distinguishing days with versus without headache (Figure 1). In contrast, the calibration was poor. The calibration plot demonstrated systematic overprediction across the range of estimated probabilities, with the smoothed calibration curve residing entirely below the 45-degree reference line. This pattern reflects miscalibration in the large, with predicted probabilities exceeding observed headache risk overall. In addition, the relationship between predicted and observed risk was nonlinear, with attenuation of observed risk at higher predicted probabilities and increasing divergence in the upper range of forecasts.

**Figure 1.**
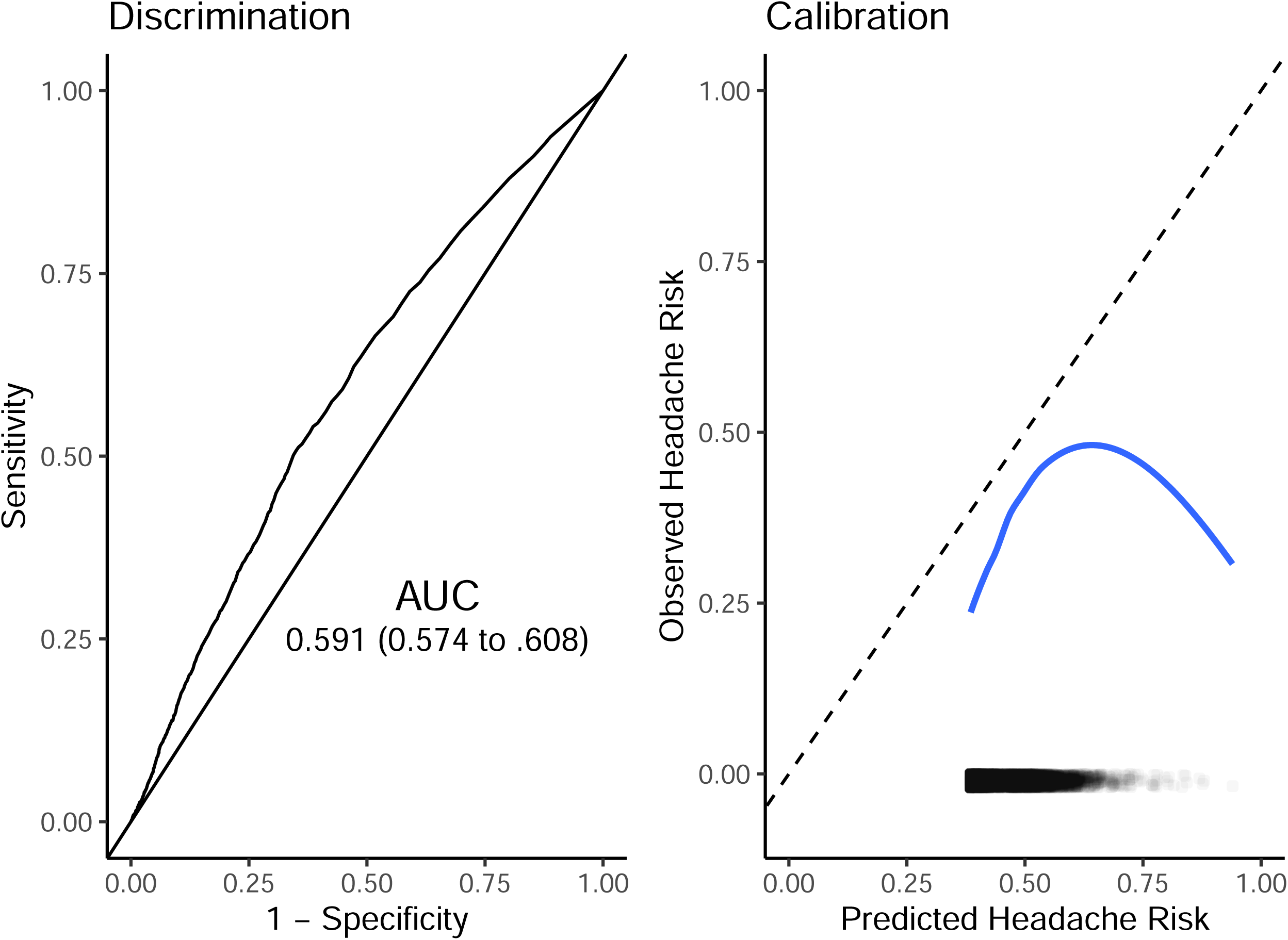
HAPRED-I external validation discrimination and calibration Receiver operating characteristic curve (left) and calibration plot (right) for all forecasts. Discrimination was modest (AUC, 0.591; 95% CI, 0.574–0.608). Calibration compares predicted and observed headache risk, with the dashed line indicating perfect agreement. The smoothed curve suggests overprediction at higher predicted risks. Rug plots show the distribution of predicted probabilities.

### Evaluation of HAPRED-II

We next evaluated how the HAPRED-II predictive performance evolved over time, with particular emphasis on whether continuous within-person updating improved the reliability of the model’s probability estimates (Figure 2). Due to the ‘warm-up’ period fewer forecasts were delivered than for the HAPRED-I model. A total of 6999 forecasts were directly delivered to participants, with headaches occurring on 2125 of 6617 evaluable forecasted days (32.1%). Early in deployment, the model exhibited poor calibration, with predicted risks deviating substantially from observed headache probabilities, consistent with the limited information available during initial days of diary collection. However, calibration improved progressively as additional participant data accrued, as the calibration curves moved steadily closer to the ideal 45-degree line across successive epochs (<14 days, 14–27 days, >27 days), indicating increasing agreement between predicted and observed headache risk across individual participants. This improvement in calibration occurred alongside increasing discrimination, with AUC increasing from 0.586 (95% CI: 0.566–0.606) during the first two weeks to 0.644 (0.615–0.670) during days 14–27 and further to 0.662 (0.628–0.695) beyond the first month. Together, these findings demonstrate that HAPRED-II’s continuous updating substantially enhances both the accuracy and reliability of its individualized forecasts over time. Additional results are reported in the Supplemental Results.

**Figure 2.**
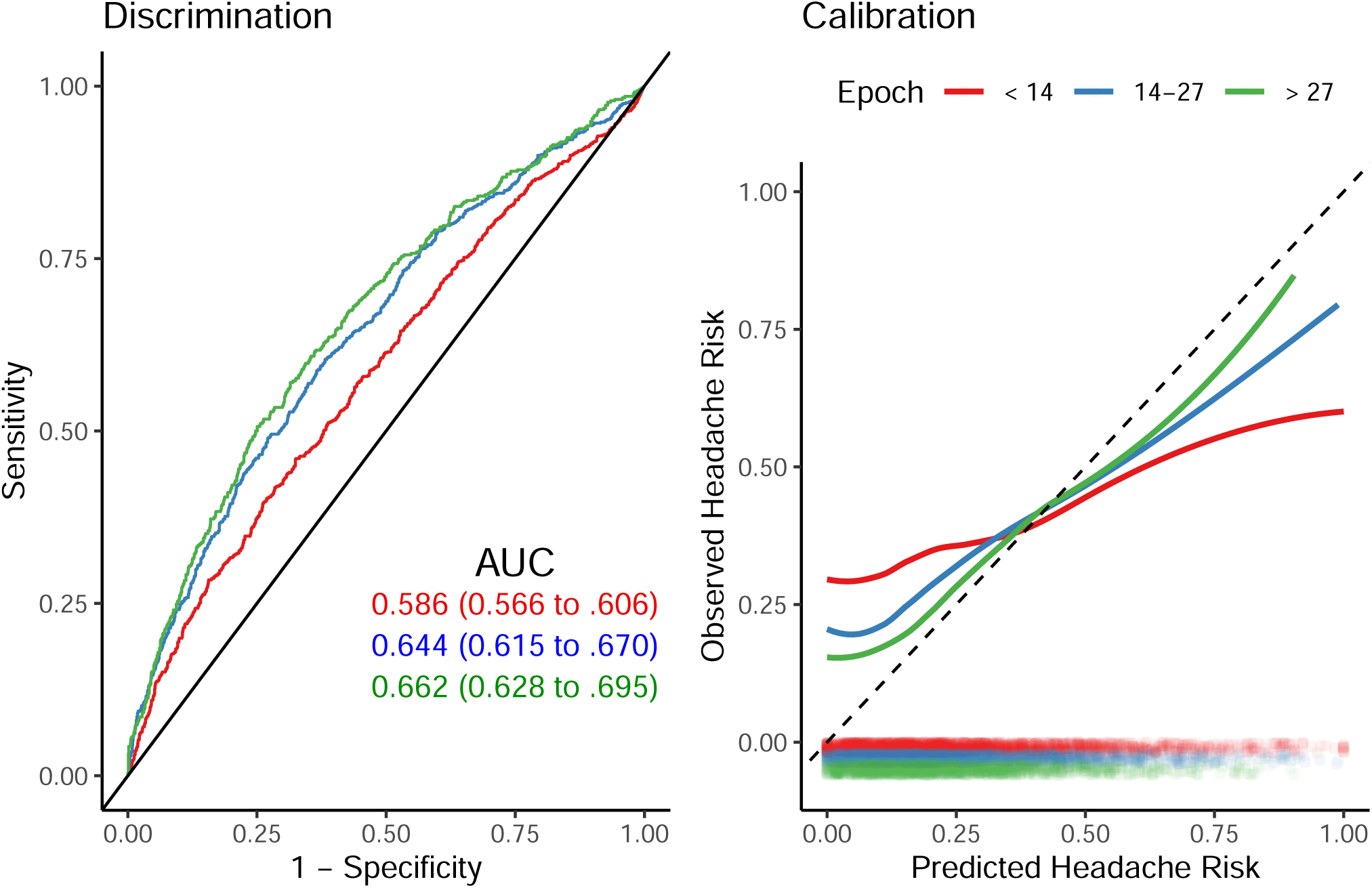
HAPRED-II dynamic discrimination and calibration Discrimination (left) and calibration (right) stratified by study epoch (<14, 14–27, >27 days). Discrimination improved over time (AUC, 0.586 to 0.662). Calibration also improved, with later epochs demonstrating closer agreement between predicted and observed risk. These findings reflect temporal updating and improved model performance with accumulating data.

### Impact of Individualized Forecasting Over Time

Figure 3 displays the evolution of smoothed daily observed headache frequency for each participant across the study period, where each gray line represents an individual’s trajectory of model-based headache risk estimates over time and the blue curve summarizes the population-average trend. Participants exhibited substantial heterogeneity in baseline headache probability at study entry; however, across follow-up there was a consistent downward shift in smoothed headache probability at the cohort level, and importantly, no overall increase in estimated headache risk was observed for participants over time. The absence of rising headache probability trajectories and the progressive decline in average estimated risk provide reassuring evidence regarding the stability and safety of deploying individualized headache risk forecasts in this population.

**Figure 3.**
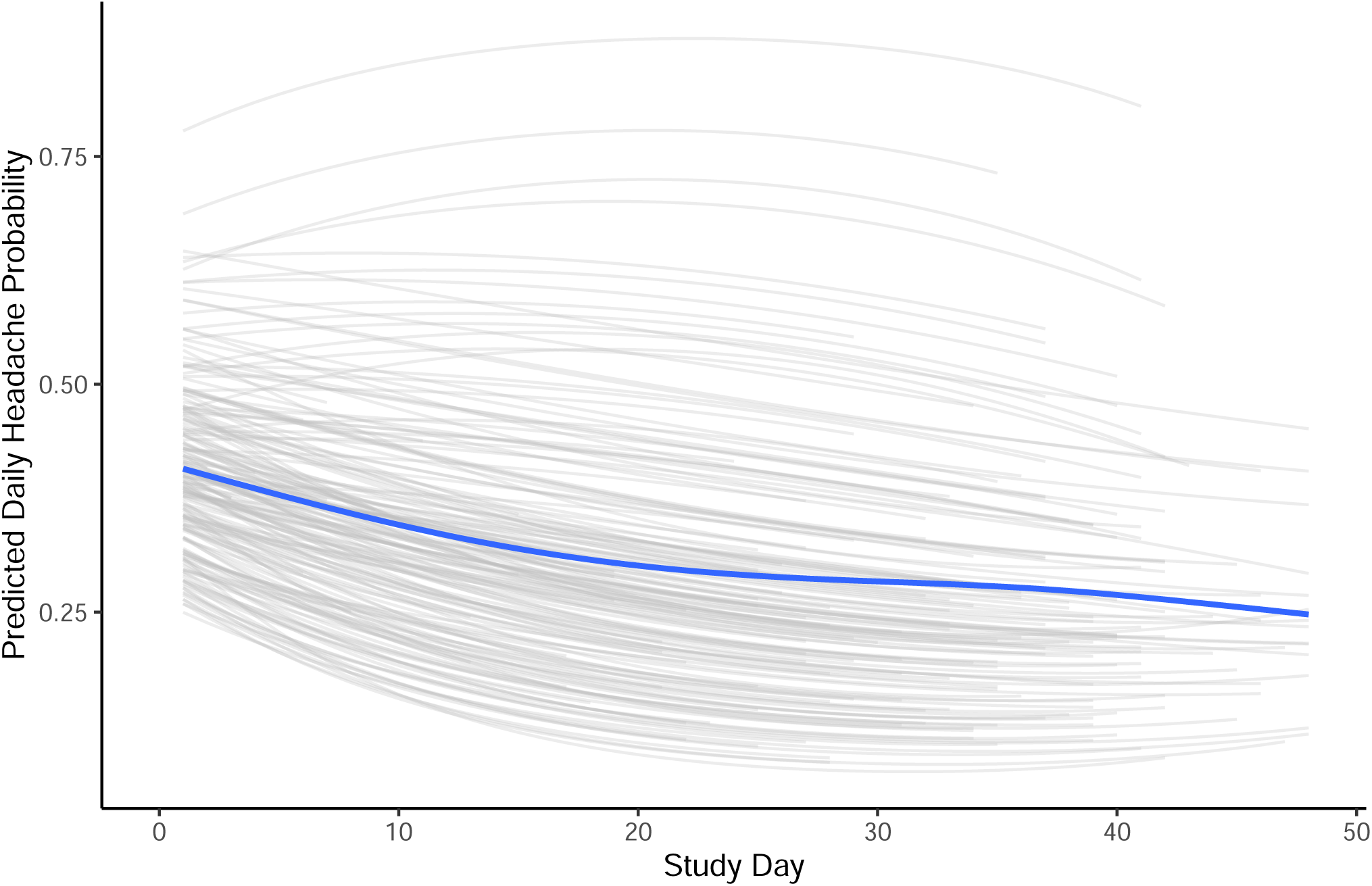
Headache frequency trajectories over time Predicted daily headache probabilities by participant over the study period. Gray lines represent individual trajectories; the blue line shows the smoothed mean. Substantial between-participant heterogeneity was observed, with a general decline in predicted risk over time, consistent with temporal updating of individualized risk estimates.

## Discussion

In this prospective external validation study of adults with recurrent headache, we conducted a strict external validation of the original HAPRED-I model.^13^ In addition to evaluating the transportability of the original model to new individuals, we implemented an updated version, HAPRED-II, that continuously learned from each participant’s accumulating data through Bayesian updating, allowing the model to become increasingly individualized over time. Using this adaptive framework, we generated and delivered daily real-time probabilistic forecasts estimating each participant’s likelihood of experiencing a headache attack within the next 24 hours. Over the course of the study, 6,999 such forecasts were delivered directly to participants, allowing us not only to evaluate model performance but also to examine the potential clinical implications of providing individualized headache risk predictions in a naturalistic setting.

We found that discrimination of HAPRED-I in an independent sample was modest, with an AUC of 0.59, slightly below the lower bound of the original internal validation estimate (AUC 0.65, 95%CI 0.60–0.67).^13^ Calibration was poor, with predicted probabilities systematically exceeding observed headache risk, particularly at higher forecasted probabilities, indicating consistent overestimation. Participants in the present study experienced lower overall headache frequency than those in the original development sample, leading to miscalibration-in-the-large and a tendency toward overprediction across the range of estimated risks. In addition, the relationship between predicted and observed risk appeared nonlinear, suggesting that the fixed coefficients derived from the development cohort did not fully capture the risk structure in this new population.

These findings are not entirely unexpected given the substantial heterogeneity that characterizes individuals with recurrent headache. Headache frequency varies markedly across patients,^23,24^ implying considerable between-person variation in baseline risk (i.e., the model intercept), and it is plausible that key predictors, such as stress, may exhibit different strengths of association with headache onset across individuals.^25^ When a single set of fixed coefficients is applied uniformly across such heterogeneous individuals, model performance may deteriorate in new samples. Taken together, these results underscore the inherent challenge of applying a single static prediction model to all individuals with recurrent headache and highlight the potential value of approaches that allow risk estimates to adapt to person-specific data over time.

Indeed, as we have observed before, allowing parameters to update within individuals substantially improves model performance over time.(see: ^19,26^) Over successive epochs of diary completion, HAPRED-II demonstrated increasing discrimination (AUC up to 0.66 beyond the first month) and calibration curves that approach the ideal 45-degree line. Although discrimination remained modest for the median individual, the improvement in calibration suggests that individualized Bayesian updating enhances the reliability of absolute risk estimates, consistent with the theoretical property that posterior predictive probabilities are calibrated with respect to the assumed data-generating model.^27,28^ Given the heterogeneity observed in participant-specific performance, adaptive modeling may be particularly advantageous in headache attack prediction which appears to be characterized by substantial within-and between-person variability.

Each evening prior to bedtime, participants received a probabilistic forecast estimating their likelihood of experiencing a headache attack during the subsequent 24 hours. Individuals were free to respond to this information in any way they chose, including modifying their activities or using medications. A potential safety concern with providing such forecasts involves the possibility of medication overuse headache. For example, a participant receiving a potentially threatening forecast (e.g., a 90% predicted chance of an attack) might preemptively take acute medication, and repeated forecast responses of this type could theoretically promote medication overuse and paradoxically increase headache frequency over time. However, we did not observe evidence consistent with this concern. Most participants experienced a reduction in headache frequency over the course of the study, and no individual demonstrated a clinically concerning increase in attacks. In addition, we did not observe increasing trajectories of predicted headache probability over time among participants receiving forecasts.

Although causal attribution is not possible in the absence of a control group, the absence of any signal suggesting worsening headache risk provides reassurance regarding the short-term safety of delivering individualized probabilistic headache forecasts directly to patients.

This study has several limitations. The cohort was predominantly female and White, which may limit the generalizability of these findings to more diverse populations that have different base rates or treatment patterns. (see: ^29^) In addition, predictors were intentionally restricted to those included in the original HAPRED-I model in order to conduct a strict external validation. While this approach preserves comparability with the original specification, it likely constrains the achievable predictive performance of the model. Primary headache attacks arise from complex and interacting physiologic, behavioral, and environmental processes,^30^ and future forecasting models may benefit from incorporating richer streams of mechanistic information such as physiologic signals, contextual exposures, or wearable-derived measures.^3,15^ In this regard, the benefits of Bayesian updating may not be fully realized until models with stronger baseline predictive performance are available. Continuous updating can help adapt predictions to an individual’s unique data-generating process, but its ultimate value depends on the quality and informativeness of the predictors entering the model. Thus, improvements in mechanistic feature sets may substantially raise the performance ceiling for adaptive forecasting systems such as in HAPRED-II.

Additional limitations relate to the clinical evaluation of the forecasting system. The impact of forecasts on medication use, functional outcomes, or quality of life was not formally assessed, and the study design did not include a control group that would permit causal inference regarding behavioral or clinical effects of receiving predictions. Finally, the outcome definition included any head pain greater than zero. Alternative definitions based on higher pain severity thresholds, attacks also exhibiting the associated symptoms of migraine, or functional impairment may yield different performance characteristics and could be explored in future work.

In conclusion, strict external validation of HAPRED-I demonstrated modest discrimination and poor calibration, underscoring the difficulty of transporting static headache forecasting models to new populations. In contrast, the continuously updating HAPRED-II framework improved both discrimination and calibration over time as it incorporated accumulating individual-level data, representing a practical implementation of real-time, personalized headache risk forecasting in a clinical research setting.

Although both models performed better than chance, their performance is probably insufficient to support most real-time clinical treatment decisions, highlighting the need for models with substantially stronger discrimination and calibration. Taken together, these findings suggest that headache forecasting is feasible when prediction systems are allowed to learn within individuals over time, but that dynamic adaptation, and likely richer predictive inputs, will be essential to achieve clinically meaningful performance.

## Supporting information

Supplemental Results

## Data Availability

All data produced in the present study are available upon reasonable request to the authors

