## Supplemental Methods for "Individualized Forecasting of Headache Attack Risk Using a Continuously Updating Model"

*Study Design and Setting*

This was a prospective, 8-week longitudinal cohort study designed for the external validation and Bayesian extension of the previously developed HAPRED-I model for forecasting daily migraine onset risk (now termed HAPRED-II). Participants were recruited from two academic medical centers: Wake Forest Health Sciences (Winston-Salem, NC) and Massachusetts General Hospital (Boston, MA). Recruitment occurred between 5/20/2015 to 1/30/2016 in Winston-Salem and 3/13/2018 to 10/18/2019 in Boston. All study activities, including screening, consent, data entry, and forecast delivery, were conducted remotely via secure web-based systems. The study was approved by the Institutional Review Boards at both participating institutions. All participants completed electronic informed consent prior to data collection.

*Participants*

Eligibility criteria were designed to closely mirror those used in the original HAPRED-I model development study,^13^ with the intention of broadly selecting community-dwelling individuals with recurrent headache who might reasonably benefit from a headache forecasting system. Adults aged 18 years or older with an International Classification of Headache Disorders, 2nd or 3rd edition (ICHD-3; ^17^) provisional diagnosis of migraine (with or without aura) or tension-type headache assessed using a structured diagnostic interview, and who reported experiencing at least one headache attack per month, were considered for enrollment. To ensure participants could successfully engage with the digital study activities, access to a computer, tablet, or smartphone and the ability to read and speak English at a sixth-grade level or higher were required. Individuals were excluded if they had a secondary headache disorder, such as one attributable to a structural neurologic condition, or if they reported a recent substantive change in headache symptoms or treatment within the past six weeks, as these factors might interfere with stable model evaluation.

*Recruitment Procedures*

Participants were recruited from the general community and from outpatient clinics affiliated with either institution. Recruitment methods included printed and digital advertisements, community fliers, local media outreach (e.g., internet, TV, print), and institutional volunteer registries. Interested individuals contacted the study team by phone or email and were provided with a brief overview of the study purpose, expectations, and voluntary nature of participation. Those expressing continued interest received a secure electronic link to review and complete the informed consent process and baseline questionnaires via REDCap.^18^

*Data collection and forecasting*

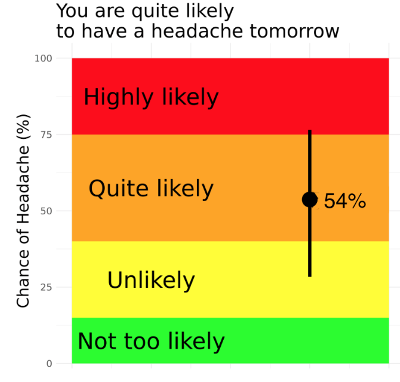
Immediately after completing baseline procedures, participants received login credentials to a secure electronic diary system. Participants completed twice-daily (morning/evening) diaries at the same time each day for 8 weeks, beginning the evening of enrollment. After completing diaries over a 7 to 14-day ‘warm-up’ period,^19^ a probabilistic forecast was delivered about the chances of experiencing a headache attack during the next 24 hours. Supplemental content 1 displays an example of a delivered forecast. At study completion, participants were invited to complete final outcome questionnaires via REDCap. Participants were compensated up to $75 based on their diary completion rate.

Individualized probabilistic forecasts were delivered following completion of the evening diary entry. Forecasts displayed the estimated probability of experiencing a headache attack during the subsequent 24-hour period, calculated as the mean of the posterior predictive distribution from the continuously updated HAPRED-II model. To communicate uncertainty, vertical bars representing the 20th and 80th percentiles of the posterior predictive distribution were displayed alongside the point estimate. Consistent with established principles of medical risk communication (see: ^31^) probabilities were also mapped onto a color-coded scale accompanied by descriptive verbal categories (e.g., “not too likely,” “unlikely,” “quite likely,” and “highly likely”). This combined numeric, visual, and verbal presentation was intended to facilitate intuitive interpretation of probabilistic risk while preserving the quantitative information provided by the model.

*Predictor Variables*

The HAPRED-I model specified only two predictor types, the degree of ‘daily hassles’ measured by the Daily Stress Inventory (DSI)^20^ and the presence-absence (binary ‘no’ or ‘yes’) of a current headache attack at the time of the diary entry, so these two variables were used to deliver forecasts in this external validation study. In the original study, the predictive performance was similar using each of the three scoring methods of the DSI (i.e., frequency of stressful events [FREQ], sum of stressful ratings [SUM], average intensity rating [AIR]),^13^  so the sum of the stress intensity ratings was used to score the DSI as this rating scheme possessed the most variance. Each of the 58 items of the DSI was rated on a 0 (‘did not occur’ to 6 (‘caused me to panic’) scale, resulting in a total SUM score that ranges from 0 to 348 points.

*Primary Outcome*

The primary outcome for model validation was the occurrence of a headache attack within the 24-hour period following each evening diary entry. Headache occurrence was defined as any diary report of head pain greater than zero, recorded in either the subsequent morning or evening diary entry of the next calendar day. This binary outcome served as the ground-truth reference for evaluating the accuracy of forecasted headache risk probabilities generated by the HAPRED-II model. All outcome data were collected prospectively through the electronic diary system, time-stamped to ensure correct temporal alignment with prediction intervals, and stored in a secure, centralized database.

*External Validation of the HAPRED-I Model*

External validation was performed by applying the previously published HAPRED-I model parameters directly to the present dataset without recalibration.

Let $Y_{i,t+1}$ represent whether participant $i$experienced a headache during the 24-hour interval following diary entry $t$. The predicted probability of headache occurrence was modeled as

$$Y_{i,t+1}\sim Bernoulli\left( p_{i,t+1} \right)$$

$$logit\left( p_{i,t+1} \right)=\alpha+\beta_{1}\left( Current_{i,t} \right)+\beta_{2}\left( Stress_{i,t} \right)$$

Where $Current_{i,t}$ denotes the presence of headache at time $t$ and $Stress_{i,t}$ denotes the Daily Stress Inventory (DSI) score. For external validation, the intercept and regression coefficients were fixed to those reported in the original HAPRED-I study $(\alpha=-0.468, \beta_{1}=0.344, \beta_{2}=0.010)$. No re-estimation, recalibration, or intercept adjustment was performed.

*HAPRED-II Individualized Prediction Model*

The HAPRED-II model extends the HAPRED-I model specification by allowing model parameters to vary across individuals.

For participant $i$, the probability of experiencing a headache attack at time $t+1$is modeled as:

$$Y_{i,t+1}\sim Bernoulli\left( p_{i,t+1} \right)$$

$$logit\left( p_{i,t+1} \right)=\alpha_{i}+\beta_{1i}\left( Current_{i,t} \right)+\beta_{2i}\left( Stress_{i,t} \right)$$

Where $\alpha_{i} now$represents the participant-specific baseline headache probability, $\beta_{1i}$represents the participant-specific effect of current headache, and $\beta_{2i}$represents the participant-specific effect of stress.

This formulation allows the model to capture heterogeneity in both baseline headache frequency and predictor effects across individuals. The real-time headache risk forecasts displayed to participants during the study were derived directly from this HAPRED-II model. These personalized forecasts were presented within each participant’s secure online diary interface at the time of each diary entry, enabling participants to view their individualized risk predictions immediately after data submission.

*Bayesian Updating Procedure*

Model parameters were estimated using Bayesian updating so that forecasts could adapt as additional participant data accumulated.

**Prior Distributions**

For each participant $i$, model parameters were assigned normal prior distributions using weak priors:

$$\alpha_{i}\sim N\left( \mu_{\alpha}= -0.909,\sigma_{\alpha}=3.3 \right)$$

$$\beta_{1i}\sim N\left( \mu_{\beta1}=0.345,\sigma_{\beta1}=2.9 \right)$$

$$\beta_{2i}\sim N\left( \mu_{\beta2}=0.013,\sigma_{\beta2}=17.7 \right)$$

**Sequential Updating**

After each new observation was recorded, posterior parameter distributions were updated according to Bayes’ rule.

Let

$$\theta_{i}=\left( \alpha_{i},\beta_{1i},\beta_{2i} \right)$$

denote the parameter vector for participant $i$. The posterior distribution after observing data up to time $t$is given by

$$p\left( \theta_{i} \mid Data_{1:t} \right)\propto p\left( Data_{t} \mid\theta_{i} \right)\times p\left( \theta_{i} \mid Data_{1:t-1} \right)$$

Posterior predictive probabilities were then used to generate forecasts for the subsequent 24-hour period:

$$p_{i,t+1}=P\left( Y_{i,t+1}=1 \mid Data_{1:t} \right)$$

This sequential updating procedure allows the model to learn participant-specific baseline risk and predictor effects over time.

*Statistical Analysis: Model Performance Evaluation*

Model performance was evaluated in terms of discrimination and calibration for both the externally validated HAPRED-I model and the continuously updated HAPRED-II model. Discrimination was quantified using the area under the receiver operating characteristic curve (AUC), which reflects the model’s ability to distinguish between days followed by a headache attack and those that were not. To account for the nesting of daily predictions within participants, a clustered bootstrap approach was used to derive 95% confidence intervals (CIs) for the AUC. In each of 1,000 bootstrap replications, participants were resampled with replacement, and the AUC was recalculated from the resampled data. Percentile-based 95% CIs were computed from the empirical bootstrap distribution of AUC values.

Calibration was assessed by plotting the observed probability of headache occurrence against the model-predicted probabilities, using locally weighted (LOESS) smoothing to visualize agreement between predicted and observed risks. For the HAPRED-I validation, predictions were generated using the fixed population intercept and slope coefficients from the original model (Formula X), without re-estimation of parameters.

For the HAPRED-II model, discrimination and calibration were examined across successive epochs of the 8-week diary period to assess improvement in predictive accuracy over time as individual-level Bayesian updating occurred. Separate ROC curves and AUC estimates were computed for three temporal segments (< 14 days, 14–27 days, and > 27 days). Confidence intervals for these epoch-specific AUCs were again derived using participant-clustered bootstrapping.

To examine heterogeneity in predictive performance, participant-specific AUCs and their corresponding 95% CIs were estimated by fitting ROC curves separately for each individual and displayed in a caterpillar plot, illustrating the range and distribution of discrimination across participants. All analyses were conducted in **R** (version 4.5.1) using the *pROC*, *boot*, *tidyverse*, and *cowplot* packages.

Sample Size Considerations

To evaluate the sufficiency of the available sample size, we used the pmsampsize^21^ approach for prediction model development with a binary outcome, specifying an anticipated C-statistic of 0.65, two predictor parameters, and an outcome prevalence of 0.287.^13^ Under these assumptions, the corresponding Cox–Snell $R^{2}$was 0.0569. The minimum required sample size was 315 participants, including approximately 91 events, which satisfied the most stringent of the three prespecified criteria implemented in pmsampsize. This sample size corresponded to a target shrinkage factor of 0.90, a margin of error of 0.05 for estimation of the model intercept, and an expected events-per-predictor parameter ratio of 45.2. Taken together, these results suggest that an independent sample of at least 315 participant-observations would be adequate to support development of the proposed binary prediction model while limiting optimism in model performance and ensuring reasonably precise estimation of the overall outcome risk.

For HAPRED-II, 230 participants contributed 8,896 evaluable forecasts. Because forecasts were clustered within individuals, the effective sample size was evaluated using a standard design effect adjustment, $N_{\text{eff}}=N/[1+(m-1)\rho]$, where $m$is the mean number of observations per participant and $\rho$is the intraclass correlation coefficient (ICC). Given an average of approximately 38.7 observations per participant, the effective sample size would fall below the required 315 only if the ICC were approximately 0.72 or greater. As such, even under implausibly high levels of within-person correlation, the available data are sufficient to support reliable evaluation of model performance.

probabilities. Report Issued for Australian Center for Risk Analysis (ACERA).
