## Supplemental Results for "Individualized Forecasting of Headache Attack Risk Using a Continuously Updating Model"

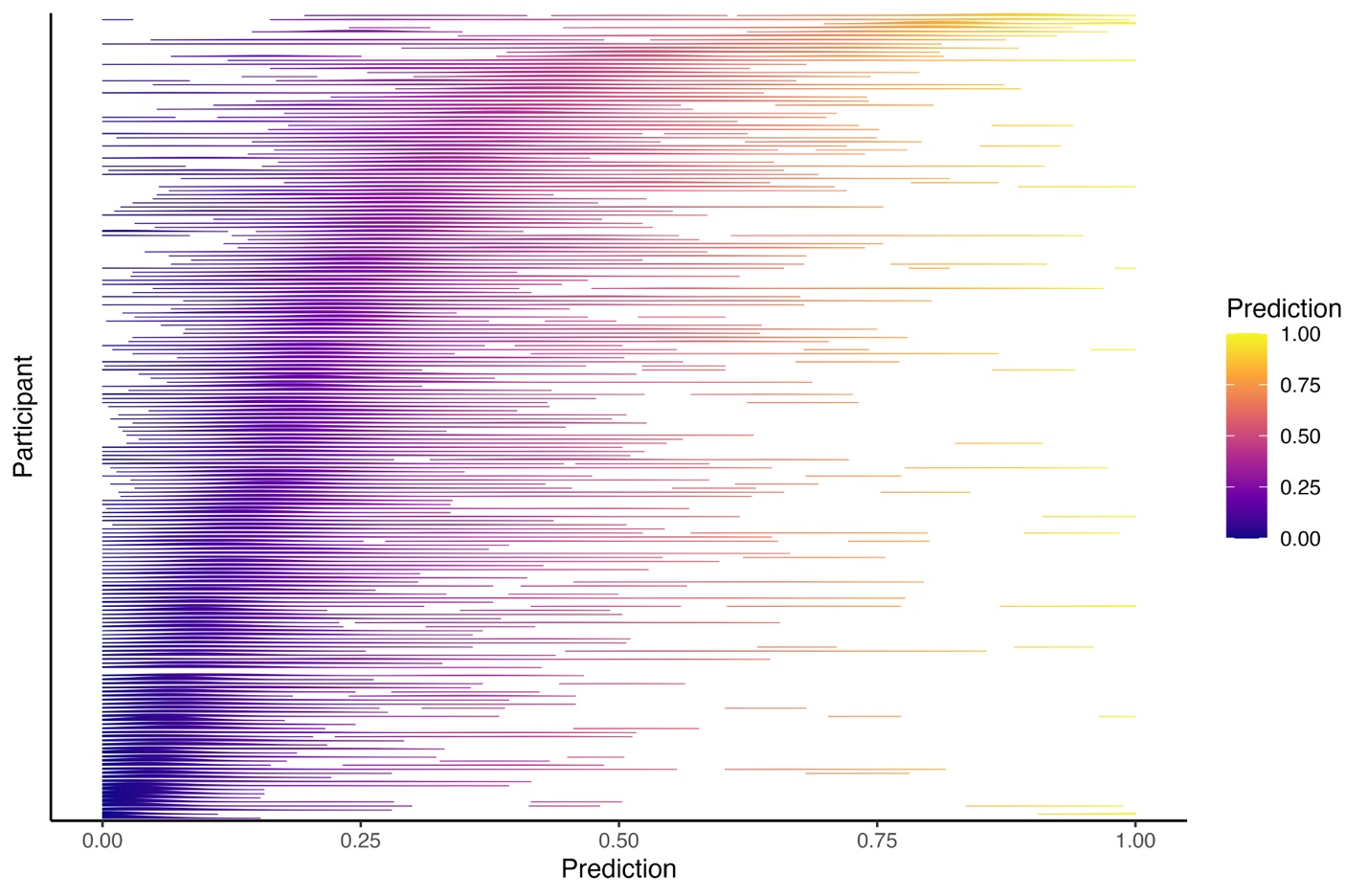

**Supplemental Figure 1** displays the distribution of predicted headache probabilities generated by the HAPRED-II model for each participant across the study period. Each horizontal line represents the distributions of an individual’s forecasts, with color indicating the predicted probability of experiencing a headache attack within the subsequent 24 hours. Participants exhibited substantial heterogeneity in their predicted risk distributions, reflecting differences in baseline headache frequency and in the strength of association between predictors and subsequent headache occurrence. Some participants showed predictions concentrated at lower probability ranges, consistent with relatively infrequent headache activity, whereas others demonstrated broader distributions extending into higher probability ranges. This heterogeneity supports the rationale for individualized model parameters and continuous within-person updating, as implemented in the HAPRED-II framework.

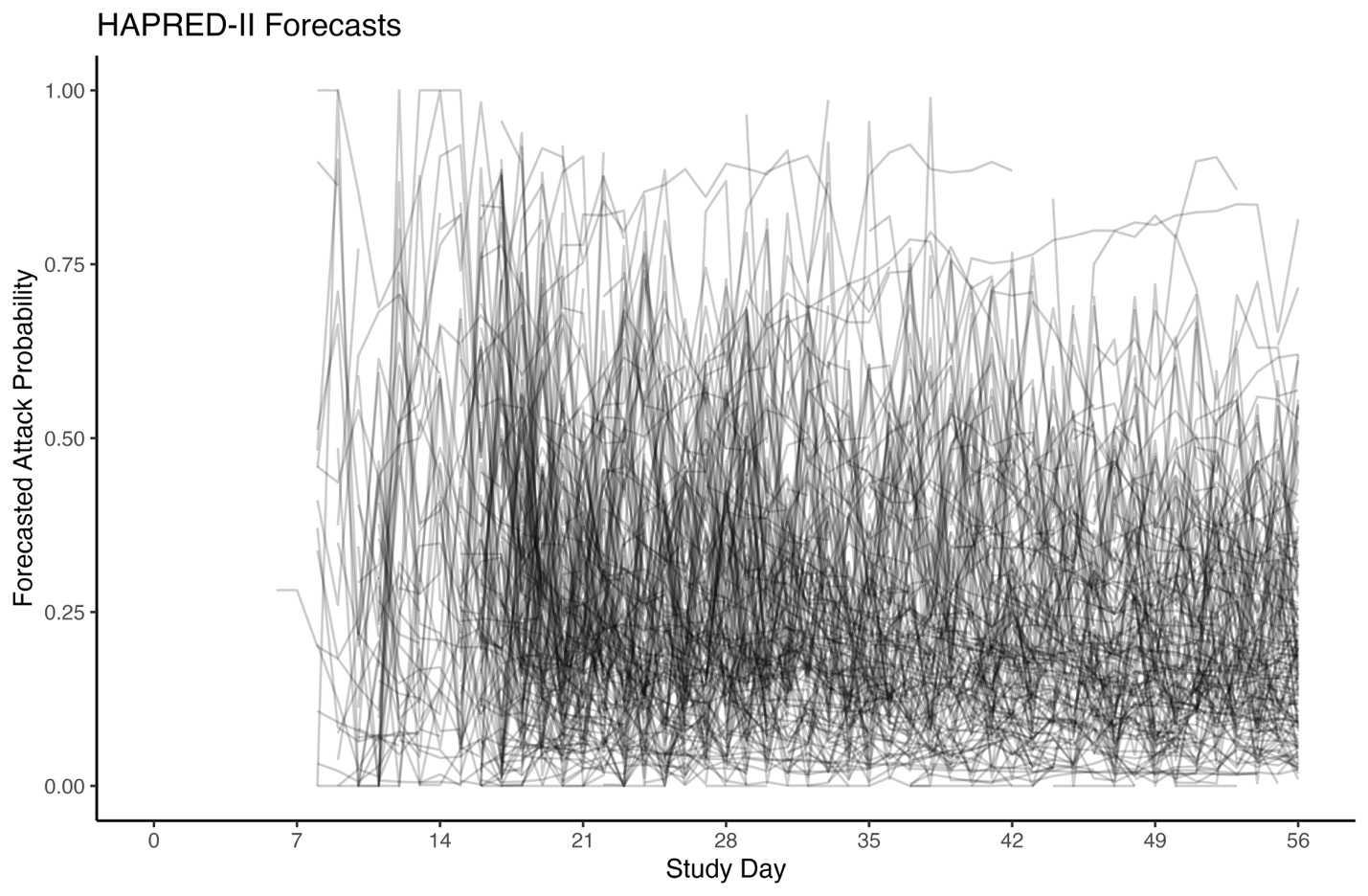

**Supplemental Figure 2** displays the sequence of individualized headache risk forecasts generated by the HAPRED-II model for each participant across the 8-week study period. Each line represents the trajectory of predicted headache probability for a single participant, with study day on the horizontal axis and the point estimate of the forecasted probability on the vertical axis. Considerable variability is evident both within and between participants, reflecting day-to-day fluctuations in predictors as well as heterogeneity in baseline headache risk across individuals. Forecast trajectories generally stabilized after the initial observation period as participant-specific parameters were updated using accumulating diary data. This visualization illustrates the dynamic nature of the HAPRED-II forecasting framework, in which predicted probabilities evolve over time as the model adapts to each participant’s observed data.

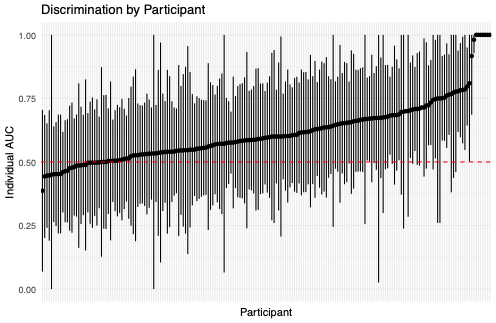

**Supplemental Figure 3** displays the discrimination performance of the HAPRED-II model at the individual participant level. Each vertical line represents the estimated area under the receiver operating characteristic curve (AUC) for a single participant, with accompanying 95% confidence intervals derived from participant-specific prediction data. Participants are ordered along the horizontal axis by their estimated AUC values. The horizontal dashed line at 0.50 represents discrimination equivalent to chance. Substantial heterogeneity in predictive performance is evident across participants, with some individuals demonstrating discrimination near chance and others exhibiting considerably higher values. This variability reflects differences in the strength and consistency of relationships between predictors and subsequent headache occurrence across individuals, further supporting the use of individualized model parameters in the HAPRED-II framework.
